# Fast initial Covid-19 response means greater caution may be needed later

**DOI:** 10.1101/2020.05.26.20112680

**Authors:** Joël J.-M. Hirschi

## Abstract

**Background:** As the Covid-19 pandemic unfolds it is becoming increasingly clear that the strength of the first wave of the epidemic varies significantly between countries. In this study a simple numerical model is used to illustrate the impact the timing of initial measures against Covid-19 has on the first wave of infection and possible implications this may have for the measures taken as the first wave is ebbing. The results highlight that delaying measures by 10 days is sufficient to largely account for the differences seen between countries such as the UK and Germany for the first wave of infections. A pronounced first wave means that a larger fraction of the total population will have been infected and is therefore likely to display immunity. Even if this fraction is far below the level needed for “herd immunity” the effective reproduction factor *R_e_* is decreased compared to a population that had no prior exposure to the virus. Even a small reduction in *R_e_* can have major influence on the evolution of the epidemic after the first wave of infections. A large first wave means the resulting value for *R_e_* will be lower than if the first wave was mild. Without either vaccine or effective treatment countries that experienced a small first wave should therefore relax measures at a slower pace than countries where the first wave was strong.

## 1. introduction

Around the Globe the Covid-19 (SARS-CoV2) pandemic is currently impacting the daily lives of billions of people. After being first reported in late 2019 in the Chinese Province of Hubei, the Covid-19 virus has spread to all continents and led to - at the time of writing - about 1/3 of the world population being subject to some form of “lockdown” rules. The paths chosen range from strict lockdowns with most people confined to their homes until the peak of the epidemic has been clearly passed (e.g. Hubei Province in China, South Korea, Italy, Spain) to more relaxed approaches where the emphasis is on minimising the impact on daily life (e.g. Sweden). Many nations are somewhere in the middle ground and have opted for various forms of “soft lockdown” where people - whilst strongly encouraged to stay at home - can leave their homes to buy groceries, to attend medical appointments or to exercise. In their approaches countries are weighing human cost and in particular the capacity of the health system to cope with a surge in the number of cases, against the cost of shutting or ramping down many sectors of the economy. Regardless of which aspect weighs more heavily on the decision-making, the adopted approaches all have in common that they want to prevent an uncontrolled, exponential increase in the number of people infected by the virus. Despite debates and in some instances tensions between scientific advice, government action and public perception there is concensus that an uncontrolled spread of the virus would come at an unacceptable human cost which would overwhelm even the best prepared and efficient health system.

All approaches that have been adopted include various forms of “social distancing” with the aim to reduce the number of direct contacts between people in order to lower the effective reproduction factor *R_e_* (i.e. the number of people an infected person will infect on average) below a value of 1. Current estimates for the reproduction factor *R_0_* in a population that had no prior exposure to Covid-19 are between 2 and 3 (e.g. Liu et al. 2020). This makes Covid-19 more contagious than for example flu viruses with *R*_0_ ≈ 1.3 but less contagious than measles with *R*_0_ ≈ 15 in previously unexposed populations. In addition to social distancing some countries also prescribe the use of face masks when people leave their home - an approach most widely seen in Asia (e.g. China, South Korea or Singapore) whereas in most western nations the choice of mask-wearing is left to the individual. Social distancing and/or wearing face masks is not a new approach but was widely used in earlier pandemics such as the Spanish flu in 1918/1919 (e.g. Martini et al. 2019; Reid et al. 2001). Such measures are simple and have been shown to be effective in slowing the spread of viruses. These low-tech measures are still the backbone of our approach to slow down pandemics. However, new technologies like contact tracing using mobile phones are increasingly being tested to localise infection hotspots and inform targeted applications of measures.

As the Covid-19 pandemic unfolds with many countries having passed or are approaching the peak of the first wave we can observe marked differences in the number of cases and deaths in countries that are broadly comparable in terms of population size, health care systems and overall living standards. For example Germany had fewer deaths in the first wave than the UK despite having a larger population 83 vs 66 millions. Whilst it will be years until the full extent of the human and economic cost of the Covid-19 pandemic is known there are already lessons that can be learned - about the initial reactions and measures in future pandemics but also what to look out for as nations gradually emerge from the first wave of a pandemic. Analytical and numerical models can be powerful tools to understand the evolution of the pandemic so far and to test possible scenarios for the evolution of the pandemic in the coming months and years (e.g. Prem et al. 2020; Kucharski et al. 2020; Tsang et al. 2020). In this article a very simple model is used to illustrate the impact the timing of measures can have on the severity of the first wave of the pandemic and the consequences of a weak or a strong first wave once the lockdown rules are gradually relaxed.

The model is introduced in section 2, the results presented and discussed in sections 3 and 4 and conclusions are given in section 5.

## 2. Method

The following introduces the model that will be used to test scenarios in which the timing for the imposition and relaxation of measures during an epidemic is varied. The basic assumptions for the model are that without imposition of any measures (i.e. daily human contacts as usual) and in a population with no prior exposure to the virus the reproduction rate *R*_0_ is assumed to be 2.5. *R*_0_ = 2.5 is at the lower end of the range of estimates for *R*_0_ for Covid-19 (e.g. Liu et al. 2020; Wu et al. 2020). An incubation time of 5 days is assumed i.e. within 5 days an infected person will on average infect 2.5 people.

The cumulative number of cases C is calculated according to

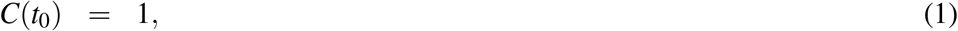

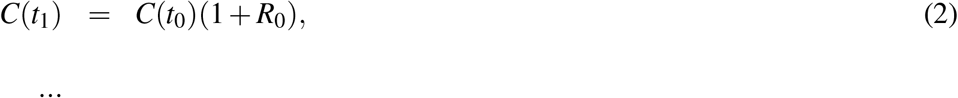

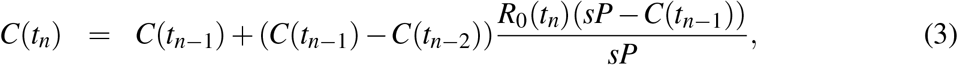

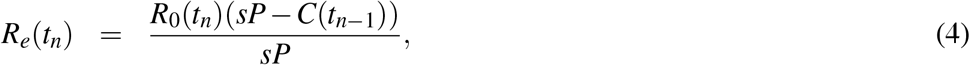

where *P* = 6.6 × 10^7^ is the total population (similar to the UK), *s =* 0.6 the fraction of the population that needs to have been infected for “herd immunity” to be reached. Since the average time between infection and first symptoms to show is about 5 days the assumption is made that Δ*t* = *t_n_* − *t_n_*_−1_ =5 days. *R*_0_ is the reproduction rate in a population that had no prior exposure to the virus. *R_e_* is the effective reproduction rate which accounts for the fact that, as the number of infections in the population rise, the ability of the virus to infect people gradually decreases. This assumes that people who had the virus will be immune to re-infection and that “herd immunity” can be achieved. A linear relationship is assumed between *C(t_n_*) and *R_e_*. More elaborate functions could be used, but for to illustrate possible types of epidemic evolutions a linear relationship is sufficient.

The reproduction rate *R*_0_*(t_n_)* as used in equation 4 can be considered as indicative of the stringency of the measures taken to slow the spread of the virus. No measures means *R_0_(t_n_) =* 2.5 and *R_0_(t_n_) =* 0 would refer to a situation where every infected person can be isolated before passing on the virus to anyone.

From *C(t_n_)* the number of new cases per day is calculated according to:

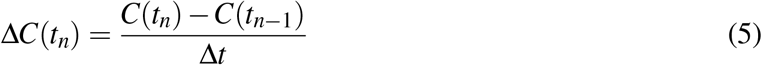

The total number of fatalities *F* at time *t_n_* is calculated according to:

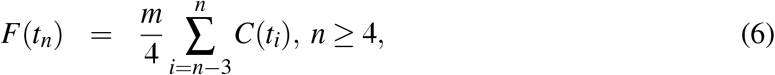

It is not yet clear what the mortality rate m for Covid-19 is. Based on the number of cases reported by the World Health Organisation fatalities make about 6.5% of the total number of cases (WHO 2020). It is estimated that the actual number of people who have been infected is at least one order of magnitude higher than the recorded number (e.g. Bendavid et al. 2020; Vardar 2020). Therefore, a mortalility rate of *m* = 0.65% is assumed here. The number of fatalities per day is:

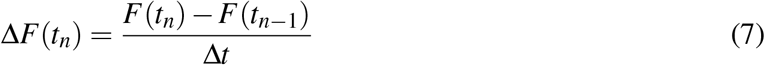

### a. Experiments

The goal of measures such as social distancing and lockdowns is to reduce *R*_0_ in order to get *R_e_* below a value of 1 before the number of infected people requiring treatment becomes too large. Figure 1 illustrates the evolution of *C(t_n_)* if no measures are taken. In this case *R_e_* solely reduces because the fraction of the population that has been infected increases. In this simple model it takes about 120 days for 60% is the population to have been infected, which here will be used as the threshold needed to reach herd immunity. From about day 80 onwards the effective reproduction rate *R_e_* starts to decrease, reaching values close to zero after about 120 days. After that time 60% of the population (about 40 Mio people) would have been infected. With the assumed mortality rate of 0.65% this would result in 250000 fatalities in a population of 66 million.

**Fig.1.**
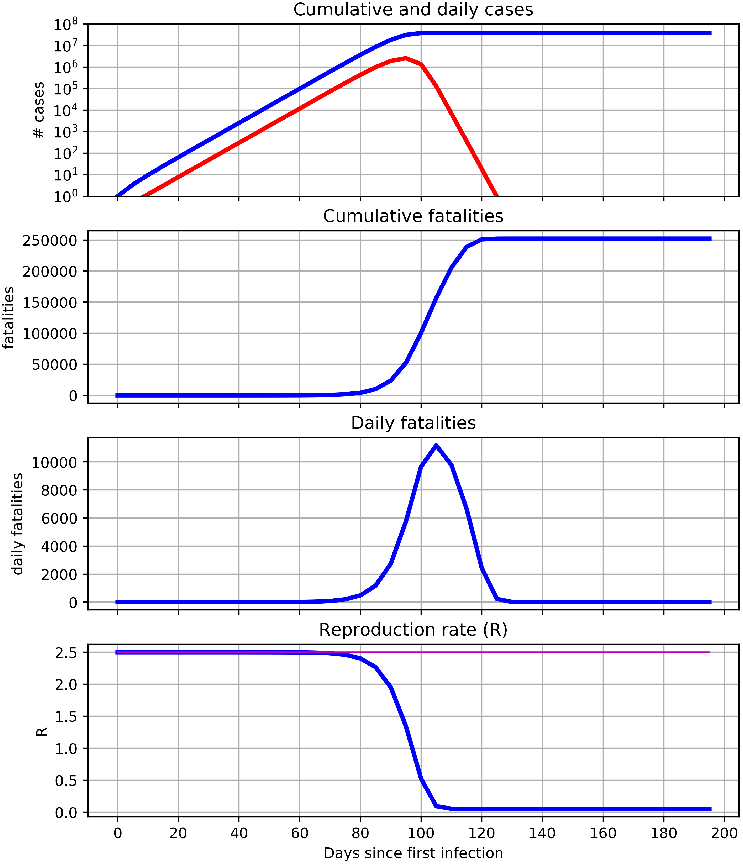
Scenario for the Covid-19 epidemic in a population of the size of the UK (66 million) assuming no measures to stop the spread of the virus are taken. Shown are: cumulative (blue) and daily (red) cases (top row), cumulative and daily fatalities (2nd and 3rd row) and the reproduction factor *R*_0_ (magenta) and “effective reproduction factor” *R_e_* (bottom)

In a set of experiments the model is used to illustrate the sensitivity of the long and short-term development of an epidemic to the timing of the initial measures (e.g. social distancing). Focus is on the first and potential second wave of infections (experiments A1, A2, B1, B2) and on possible long-term evolutions of the epidemic during the years following the first infection (C1, C2, D1, D2). The details for the experiments are listed in table 1. All experiments have in common that the first measures (reducing *R*_0_ from 2.5 to 1.1) start either on day 75 (A1, B1, C1, D1) or 65 (A2, B2, C2, D2). “Lockdown” measures which reduce *R*_0_ from 1.1 to 0.85 start on day 80 for all experiments. The experiments (A, B, C, D) then differ on the timing and type of measures following the lockdown on day 80. All experiments are summarised in table 1.

**Table 1.** Experiment overview. Measures start either on day 75 or 65 and lockdown starts on day 80 in all cases. Lockdown is either relaxed on day 115 (A1, A2, C1, C2, D1, D2 - *R_0_ =* 1.1 or 1.05) or completely relaxed (*R*_0_ *=* 2.5) if there have been no new daily new cases for 50 days (B1, B2). Further relaxation is either on day 165 (A1, A2); if daily fatalities Δ*F <* 50 (C1, C2); or if the effective reproduction rate *R_e_ <* 1 (D1, D2). Measures are tightened again on day 185 (A1, A2 - *R_0_ =* 1.1) or if the daily fatalities Δ*F* > 75 (C1, C2 - *R*_0_ = 1.05)

| Experiment | Start measures<br>[day] | Lockdown<br>[day] | Relax measures<br>[day, $\Delta C$] | Relax further<br>[day, $\Delta F$, $R_e$] | Tighten again<br>[day, $\Delta F$] |
| --- | --- | --- | --- | --- | --- |
| A1 | 75 ($R_0 = 1.1$) | 80 ($R_0 = 0.85$) | 115 ($R_0 = 1.1$) | 165 ($R_0 = 1.7$) | 185 ($R_0 = 1.1$) |
| A2 | 65 ($R_0 = 1.1$) | ” | ” | ” | ” |
| B1 | 75 ($R_0 = 1.1$) | ” | If $\sum_{i=n-10}^n \Delta C(t_i) = 0$: $R_0 = 2.5$ | — | — |
| B2 | 65 ($R_0 = 1.1$) | ” | ” | — | — |
| C1 | 75 ($R_0 = 1.1$) | ” | 115 ($R_0 = 1.05$) | If $\Delta F < 50$: $R_0 = 1.4$ | If $\Delta F > 75$: $R_0 = 1.05$ |
| C2 | 65 ($R_0 = 1.1$) | ” | ” | ” | ” |
| D1 | 75 ($R_0 = 1.1$) | ” | ” | If $R_e(t_n) < 1$:<br>$R_0(t_{n+1}) = R_0(t_n) + (1 - R_e(t_n))$ | — |
| D2 | 65 ($R_0 = 1.1$) | ” | ” | ” | — |

To illustrate that the model can simulate realistic evolutions of the first wave of the Covid-19 epidemic the simulated number of fatalities is aligned with the numbers recorded in the UK and Germany. Model and observations are temporally aligned from the time onward when UK, German, and simulated total numbers of fatalities first reach or exceed 10. In the model this threshold is reached after 60 days. Data for the UK and Germany are available from Worldometers (2020).

## 3. Results

The results shown in Figures 2 and 3 highlight the influence of timing of the first measures on the severity of the first wave of an epidemic. A difference of 10 days in the starting time of initial measures which reduce *R*_0_ from 2.5 to 1.1 (soft lockdown) is sufficient for the model to simulate evolutions of the number of deaths and death rate during the first wave which are comparable to the numbers recorded in the UK and Germany. If first measures are introduced on day 75 (experiments A1, B1, C1, D1) the maximum daily fatalities reach a maximum of just over 1000 deaths a day and by day 115 the cumulative fatalities exceed 30000. In contrast if first measures start on day 65 (experiments A2, B2, C2, D2) the model simulates a peak in daily fatalities of just under 250 and a total number of fatalities of under 8000 - even though the lockdown starts on day 80 (*R*_0_ = 0.85) in all experiments.

**Fig. 2.**
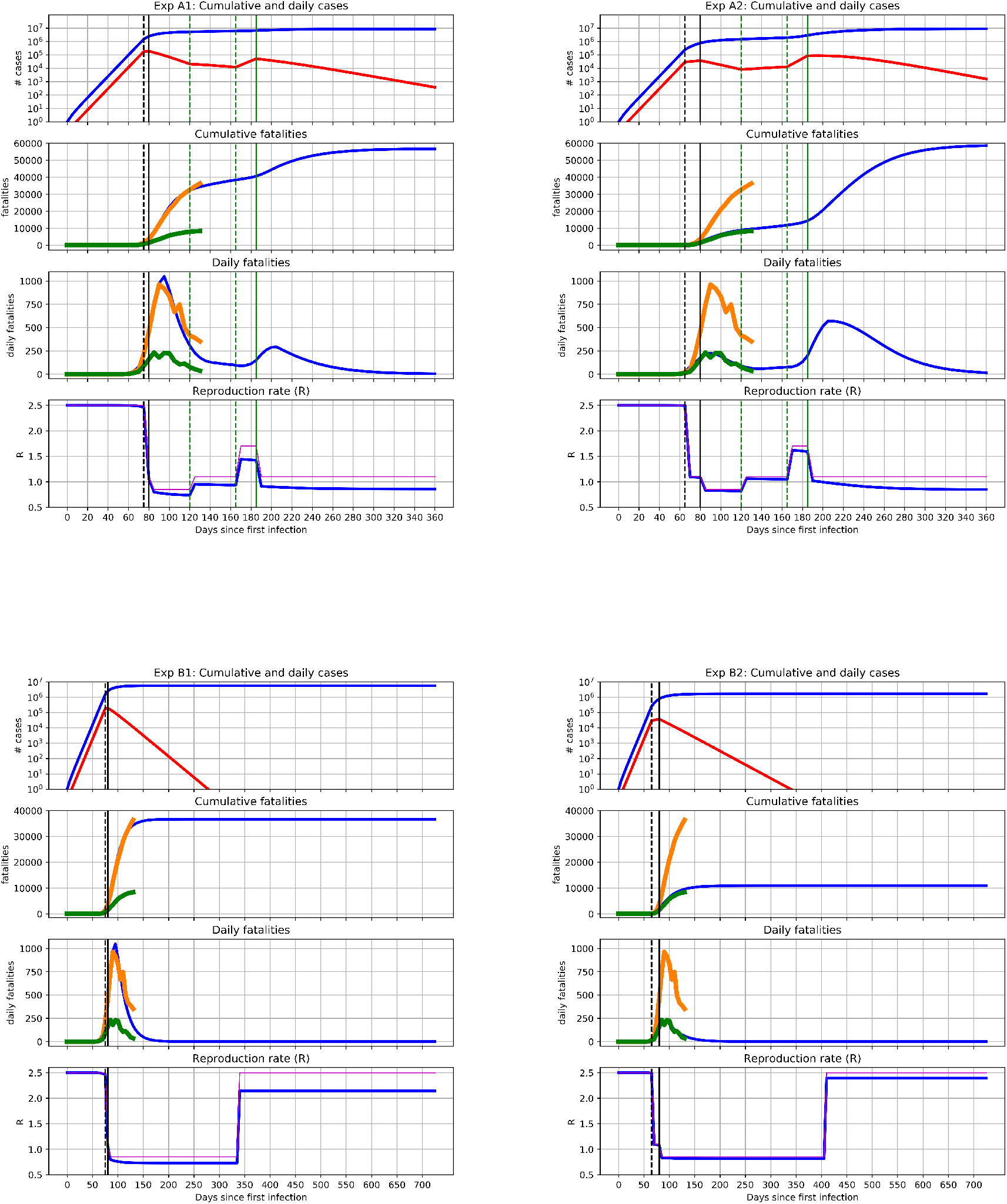
As Figure 1 for experiments A1, A2, B1, B2 (see Table 1 for details). Vertical lines show the timing when different measures were taken: **black dashed** - begin of measures (steps in the values of *R*_0_ (magenta) and *R_e_* (blue)), **solid black** - Lockdown, **green dashed** - first and second easing of lockdown, solid green - 2nd lockdown. For the first wave the model is compared to the fatalities reported in the UK (**orange line**) and Germany (**green line**).

**Fig. 3.**
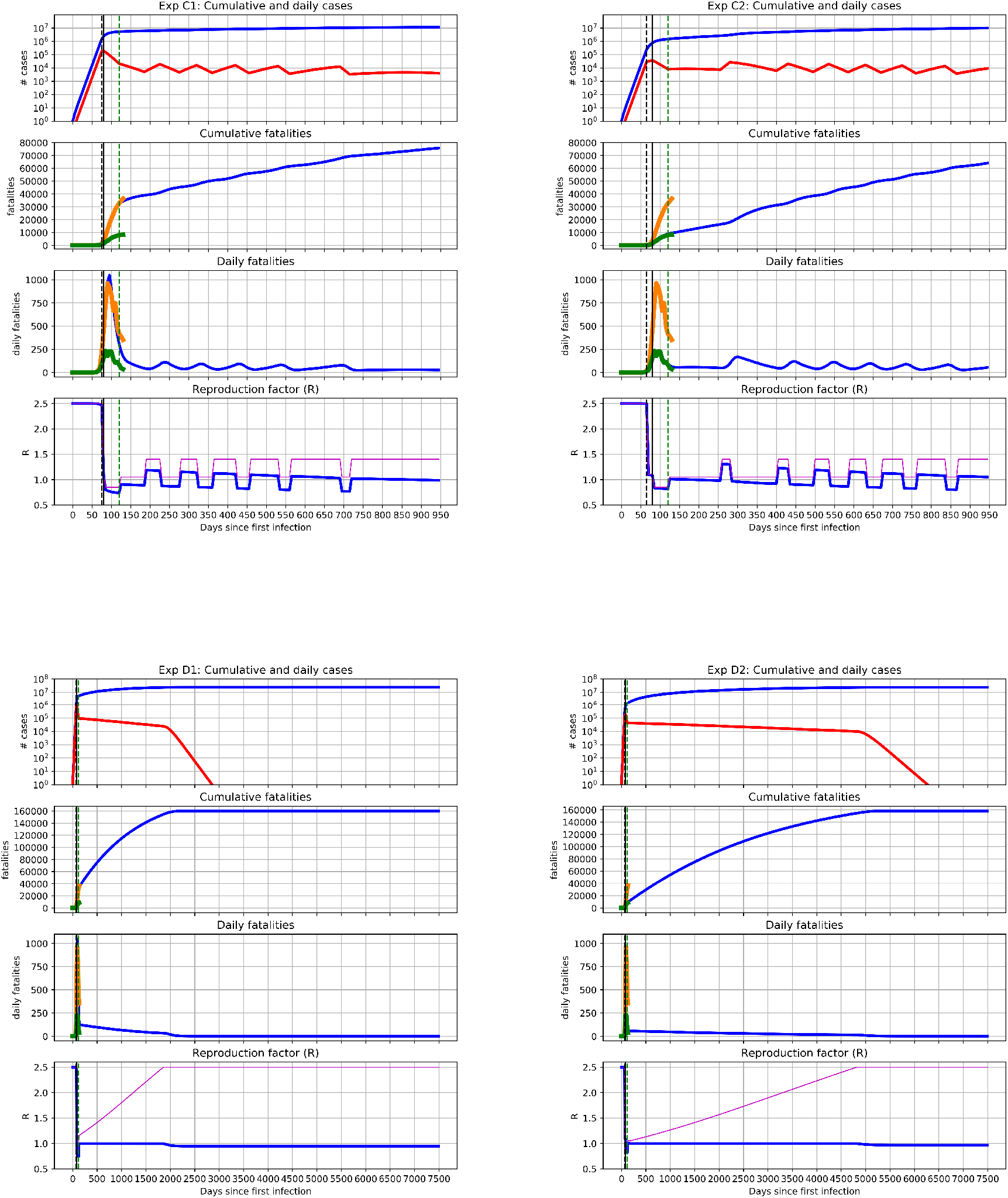
As Figure 2 for experiments C1, C2, D1, D2 (see Table 1 for details). Vertical lines show the timing when different measures were taken: **black dashed** - begin of measures (steps in the values of *R*_0_ (magenta) and *R_e_* (blue)), **solid black** - Lockdown, **green dashed**. For the first wave the model is compared to the fatalities reported in the UK (**orange line**) and Germany (**green line**).

Except for experiments B1 and B2 where a long lockdown allows the number of cases to fall to zero, the lockdown measures are relaxed on day 115. In experiments A1 and A2 there is a further relaxing of measures on day 165 which allows a second wave to develop. This second wave is much more pronounced for experiment A2 (”German” case) than for experiment A1 (”UK” case). The respective peaks for daily fatalities are about 750 and 300. By day 280 a similar number of cumulative fatalities (≈ 60000) is reached in both cases. The reason for the difference in the amplitude of the second wave is the effective transmission rate *R_e_*. In A1 the simulated total number of cases *C* is just under 6 milliom people by the end of the lockdown (i.e. just under 10% of the total population). However, less than two million people (around 3% of the total population) would have been infected by the end of the lockdown in A2. These numbers are in the same order as studies suggest for St Clara County in California (Bendavid et al. 2020) or Spain (Vardar 2020). This difference is significant when it comes to the effective transmission rate *R_e_*: In neither of the two cases do we get close to the 60% level needed for herd immunity, however, the partial herd immunity is more pronounced in A1 than A2. When considering measures that aim at getting *R_e_* close to or ideally below a value of 1 having 3% or 9% of the population who went through the infection can be enough to get an effective reproduction rate which is just over or just under the critical threshold of 1. This can be clearly seen for experiments A1 and A2 (Figure 2): After the lockdown ends on day 115, *R_e_* is below 1 for experiment A1 until a further relaxation allows *R*_0_ and *R_e_* to increase to 1.7 and about 1.4 in A1 whereas *R_e_* increases to about 1.6 in A2 leading to a markedly stronger second wave. As mentioned in section 2 *R*_0_ does change according to the assumed stringency of measures.

Experiments B1 and B2 (Figure 2) illustrate the evolution for a prolonged lockdown where *R*_0_ = 0.85 until the number of daily cases Δ*C* decreases to zero. This point is reached after about 280 days in B1 and 340 days in B2. The numbers of fatalities plateau at about 37000 and 10000 cases for B1 and B2, respectively.

Experiments C1 and C2 show the possible evolution for repeated loosening and tightening of the measures if daily fatalities Δ*F* < 50 and Δ*F* > 75. Rather than a second wave the evolution is characterised by a number of “ripples” as measures are loosed or tightened. The number of fatalities gradually increases in both C1 and C2 but the slope flattens as the value of *R_e_* decreases. This is more pronounced in C1: the stronger first wave leads to a larger difference between *R*_0_ and *R_e_*. As mentioned earlier the reproduction rate *R*_0_ can be regarded as a measure for how stringent measures are: *R*_0_ = 2.5 means that daily life is not restricted in any way (as shown in Figure 1 this comes at a high human cost) and *R*_0_ = 0 means that there are no new infections. This is only possible if every infected person is completely isolated and is unrealistic once the epidemic is spreading through the population. Experiments D1 and D2 illustrate the case where *R*_0_ is gradually allowed to increase starting from *R*_0_ = 0.85 during the lockdown to *R*_0_ = 2.5. The rate of increase in *R*_0_ is determined by how large the tendency of *R_e_* to fall is. The large first wave in D1 means *R*_0_ can increase more quickly than in D2. In D1 *R*_0_ = 2.5 after about 5 years whereas it takes about 13 years in D2. For both D1 and D2 the number of fatalities is about 160000.

## 4. Discussion

The cases shown in Figures 2 and 3 are very idealised. There is little doubt that if A2 (”German” case - Figure 2) were to start developing in the real world, measures would be tightened again before day 185 therefore avoiding the pronounced second wave. Given that *F* and Δ*F* are recorded, authorities would be aware of this development and could act accordingly. Experiments B1 and B2 have the lowest number of fatalities but they would require a long lockdown. With lockdowns now being gradually eased this is not the path that most countries have chosen.

The scenarios C1 and C2 Figure 3 are more realistic as the extent of measures can adjust to changes in the number of recorded infections and deaths. Here we can see that after the first lockdown there are several phases where measures are tightened or loosened for short periods resulting in “ripples” in the number of infections and fatalities but avoiding a strong second wave. The thresholds of daily fatalities used to tighten or loosen measures are identical in C1 and C2. However, the effective reproduction factor *R_e_* is consistently lower in C1 than in C2. In order to maintain the advantage of a clearly lower number of fatalities during the first wave the measures in C2 would need to be slightly more stringent than in C1 i.e. *R*_0_ would need to be consistently lower. However, if one assumes that a vaccine (or an effective treatment) becomes available within 1 to 1.5 years C2 leads to a better outcome than C1. However, C1 would allow a daily life that is closer to “normal” than C2. By specifically targeting “hotspots” where infections flare up again it may be possible to consistently maintain an overall value of *R_e_* < 1 without the need to tighten measures everywhere (the latter is the underlying assumption in all experiments). This may lead to evolutions of cases and fatalities which fall somewhere between experiments B1/2 and C1/2.

The scenarios illustrated in Figures 2 and 3 show that both the timing of the first measures as well as the choices made once the first wave is ebbing are crucial. Early action is the most effective way to reduce the amplitude of a first wave. Even delaying measures by a few days can result in many more lives being lost in that first wave. During the Covid-19 pandemic some governments chose not to act on initial warnings but only once it became obvious that the pandemic had taken a firm foothold. Here, it is also important to acknowledge the difficulty in knowing how far into a wave of infection a country actually is. Even in neighbouring countries the wave may be at an earlier or later stage and taking identical measures at the same time may lead to very different outcomes. If by the time the warnings come in the epidemic is more advanced than expected even swift action will not be sufficient to avoid a major first wave. On the other hand if the development of the wave lags expectations late action may still be sufficient to largely suppress the first wave. The large uncertainty regarding the stage of an epidemic means that a good (or bad) outcome can also be down to luck.

However, luck can no longer be a factor after the first wave. Assuming that the mortality rate and level of health care are broadly similar in different countries a large number of deaths in proportion to the total population is a strong indicator that a higher percentage of the population has been infected than if the number of deaths is low. The results shown in Figures 2 and 3 suggest that when gradually easing the lockdown, countries which had a mild first wave (e.g. Germany, Austria, Norway, South Korea) may opt for a slightly slower unwinding of measures than countries that experienced a major first wave. Experiencing a major first wave means that there is likely to be a higher “partial herd immunity” and that an effective reproduction rate *R_e_* < 1 can be achieved with more relaxed measures than in countries where the partial herd immunity is lower. This effect can already be significant even if less than 10% of the total population have been infected (e.g. Figure 2). Current studies suggest that only 10% or less of the total number of infections have been detected and that less than 10% of the population has been infected (Bendavid et al. 2020; Vardar 2020).

At this point it is worth remembering that if one assumes a virus against which a vaccine is years away it is likely that the majority of the population will eventually be infected. Furthermore, “herd immunity” can only be achieved if people who had the infection develop long-term immunity. Whether this is the case is still subject of ongoing research (e.g. Prompetchara et al. 2020; Shi et al. 2020; Grifoni et al. 2020; Braun et al. 2020). The only time when eradication of a virus is possible without a vaccine is at the very onset of the outbreak provided the outbreak is localised and that infected people can be isolated until they are no longer infectious. However, the window of opportunity for this is short and by the time health services and authorities become aware of (or acknowledge) the situation it may already be too late. Since people with Covid-19 can be infectious before showing any symptoms (or indeed without developing systems) (e.g. Cascella et al. 2020; Tindale et al. 2020) and that much international travel carried on as normal in the early stages of the pandemic, containment was always going to be difficult.

## 5. Conclusions

A simple model has been used to simulate different scenarios for the Covid-19 epidemic in a population of the size of the UK. The findings suggest that:

- Timing of inital measures is key not just for the first wave of an epidemic but - in the absence of either a vaccine or effective treatment - also for the longer term evolutions for years following the initial outbreak.
- Shifting the implementation time of first measures by 10 days could explain differences in the number of recorded fatalities during the Covid-19 pandemic in countries such as the UK or Germany.
- A large first wave means that easing of lockdown measures can occur faster than if the first wave was small. The reason for this is a lower effective reproduction factor after a strong first wave due to partial “herd immunity”.
- Relaxing measures too much after a small first wave risks cancelling out the advantage gained by the timely initial response to the pandemic.
- Whether intial measures can largely suppress a the first wave of an epidemic is a combination of timely action and heeding advice as well as luck as at the time when first measures are taken it is difficult to know how far into the epidemic a country/region/town has progressed.

## Data Availability

The data used in this study are freely available from Worldometers.com

## Acknowledgments

Comments by Bablu Sinha are gratefully acknowledged. This research received no specific grant from any funding agency, commercial or not-for-profit sectors.

## References

Bendavid, E., and Coauthors, 2020: COVID-19 Antibody Seroprevalence in Santa Clara County, California. *MedRxiv*.

Braun, J., and Coauthors, 2020: Presence of sars-cov-2 reactive t cells in covid-19 patients and healthy donors. *medRxiv*, doi:10.1101/2020.04.17.20061440, URL https://www.medrxiv.org/content/early/2020/04/22/2020.04.17.20061440, https://www.medrxiv.org/content/early/2020/04/22/2020.04.17.20061440.full.pdf.

Cascella, M., M. Rajnik, A. Cuomo, S. C. Dulebohn, and R. Di Napoli, 2020: Features, evaluation and treatment coronavirus (COVID-19). Statpearls [internet], StatPearls Publishing.

Grifoni, A., and Coauthors, 2020: Targets of T cell responses to SARS-CoV-2 coronavirus in humans with COVID-19 disease and unexposed individuals. Cell, https://doi.org/10.1016/j.cell.2020.05.015.

Kucharski, A. J., and Coauthors, 2020: Early dynamics of transmission and control of covid-19: a mathematical modelling study. The lancet infectious diseases.

Liu, Y., A. A. Gayle, A. Wilder-Smith, and J. Rocklöv, 2020: The reproductive number of COVID-19 is higher compared to SARS coronavirus. Journal of travel medicine.

Martini, M., V. Gazzaniga, N. L. Bragazzi, and I. Barberis, 2019: The Spanish Influenza Pandemic: a lesson from history 100 years after 1918. Journal of Preventive Medicine and Hygiene, 60 (1), E64.

Prem, K., and Coauthors, 2020: The effect of control strategies to reduce social mixing on outcomes of the covid-19 epidemic in wuhan, china: a modelling study. The Lancet Public Health.

Prompetchara, E., C. Ketloy, and T. Palaga, 2020: Immune responses in COVID-19 and potential vaccines: Lessons learned from SARS and MERS epidemic. Asian Pac J Allergy Immunol, 38 (1), 1–9.

Reid, A. H., J. K. Taubenberger, and T. G. Fanning, 2001: The 1918 Spanish influenza: integrating history and biology. Microbes and infection, 3 (1), 81–87.

Shi, Y., and Coauthors, 2020: Covid-19 infection: the perspectives on immune responses. Nature Publishing Group.

Tindale, L., and Coauthors, 2020: Transmission interval estimates suggest pre-symptomatic spread of COVID-19. *MedRxiv*.

Tsang, T. K., P. Wu, Y. Lin, E. H. Lau, G. M. Leung, and B. J. Cowling, 2020: Effect of changing case definitions for covid-19 on the epidemic curve and transmission parameters in mainland china: a modelling study. The Lancet Public Health.

Vardar, S., 2020: Estudio de seroprevalencia: sólo el 5% de los espaóoles tiene antic-uerpos frente al coronavirus. URL https://www.elmundo.es/ciencia-y-salud/salud/2020/05/13/5ebc25ae21efa09e608b45f5.html, 13 May 2020, El Mundo.

WHO, 2020: WHO Coronavirus Disease (COVID-19) Dashboard. URL https://covid19.who.int, World Health Organisation.

Worldometers, 2020: Coronavirus. URL https://www.worldometers.info/coronavirus/country/uk/, https://www.worldometers.info/coronavirus/country/germany/.

Wu, J. T., K. Leung, and G. M. Leung, 2020: Nowcasting and forecasting the potential domestic and international spread of the 2019-ncov outbreak originating in wuhan, china: a modelling study. The Lancet, 395 (10225), 689–697.

